# Genomic surveillance reveals age-structured SARS-CoV-2 transmission across demographics and settings

**DOI:** 10.1101/2025.04.04.25324273

**Authors:** Gage K. Moreno, Taylor Brock-Fisher, Lydia A. Krasilnikova, Stephen F. Schaffner, Meagan Burns, Carolyn E. Casiello, Katelyn S. Messer, Brittany Petros, Ivan Specht, Katherine C. DeRuff, Katherine J. Siddle, Christine Loreth, Nicholas A. Fitzgerald, Heather M. Rooke, Stacey B. Gabriel, Sandra Smole, Shirlee Wohl, Daniel J. Park, Lawrence C. Madoff, Catherine M. Brown, Bronwyn L. MacInnis, Pardis C. Sabeti

## Abstract

Understanding respiratory virus transmission requires integrating pathogen genomic data with detailed epidemiological context. We analyzed >85,000 SARS-CoV-2 genomes linked to epidemiological data collected in Massachusetts from November 2021 to January 2023.

Collected in 666 settings, including schools, colleges, and skilled nursing facilities, this dataset enabled direct comparison of within-setting transmission to community-wide risk. Elevated transmission was concentrated in specific age-defined subpopulations–older adolescents in schools, undergraduates in colleges, and residents in nursing facilities–with little additional risk among other groups, notably including staff. Young adults, particularly in colleges, played a disproportionate role in spread, and new viral lineages consistently expanded first within this group. Viral spread followed reproducible urban-to-rural gradients, and vaccination was associated with reduced onward transmission. We further estimated sequencing thresholds required for timely detection of emerging variants. Together, these findings reveal consistent demographic and spatial structure in SARS-CoV-2 transmission that can inform future viral respiratory pathogen genomic surveillance.

## INTRODUCTION

Genomic surveillance transformed understanding of SARS-CoV-2 evolution and spread during the COVID-19 pandemic. Most large-scale surveillance efforts were designed primarily to detect and track emerging variants of concern, often with limited linked epidemiological context. In contrast, studies integrating viral genome data with detailed metadata have enabled more comprehensive analyses of the pandemic, yielding important insights into risk factors for infection^1,2,3^ and outbreak dynamics^4–7^. Nevertheless, many questions remain about the epidemiological drivers that shaped SARS-CoV-2 transmission, and addressing these questions using linked genomic and epidemiological data is likely to yield broadly relevant insights into the spread of respiratory viruses.

The Broad Institute’s clinical laboratory provided large-scale, PCR-based COVID-19 diagnostic testing throughout the acute phase of the pandemic, with a focus in its home state of Massachusetts, USA. The laboratory served public testing programs as well as educational, medical, congregate living, and other settings. The Broad Institute also sequenced a subset of positive samples through the US CDC’s National SARS-CoV-2 Strain Surveillance (NS3) program. By early 2023, these efforts had generated more than 37 million diagnostic tests and 175,000 SARS-CoV-2 genomes, representing ∼5% of all tests and genomes produced in the United States.^8,9^ In parallel, the Massachusetts Department of Public Health (MDPH) and municipal public health departments collected extensive epidemiological data on COVID-19 cases statewide. Combining these efforts, we assembled a dataset of over 130,000 SARS-CoV-2 genomes collected in Massachusetts from late 2021 through early 2023, spanning the late Delta wave and successive Omicron subvariant waves. Over 85,000 genomes were linked to epidemiological data, including age, gender, vaccination status, municipality of residence, and testing setting.

This dataset enabled unusually detailed analyses of SARS-CoV-2 transmission across institutional and community environments. Metadata describing the testing setting, including schools, colleges, and skilled nursing facilities (SNFs), allowed systematic comparison of transmission dynamics across varied settings and among sub-populations within them.

Municipality-level residence information provided fine-scale geographic resolution. Demographic and vaccination data, available for a substantial fraction of cases, added further individual-level context. Together, these features created an opportunity to examine how age, setting, geography, and vaccination status shaped transmission patterns at scale.

In this study, we integrated genomic, geospatial, and epidemiological data to examine how SARS-CoV-2 transmission varied across age groups, institutional environments, and viral lineages. We analyzed how new variants were introduced and propagated, how transmission risk differed across demographic and contextual settings, and how these dynamics shifted across epidemic waves. We also assessed associations between vaccination status and both infection risk and onward transmission, and estimated the sampling intensity required for timely detection of emerging lineages. Together, these analyses define demographic and spatial structure in viral spread and provide insights to guide future public health genomic surveillance and response strategies for viral respiratory pathogens.

## RESULTS

### Over 85,000 SARS-CoV-2 genomes paired to individualized epidemiological data

We analyzed 134,785 high-quality (≥80% complete) SARS-CoV-2 genomes generated by the Broad Institute from residual positive diagnostic samples collected from Massachusetts residents between November 1, 2021, and January 17, 2023. These genomes comprised 492 named Pango lineages, which fell into seven dominant lineages: Delta (B.1.1.617.2*, where the asterisk indicates the inclusion of sublineages^10^) and the BA.1*, BA.2*, BA.4*, BA.5*, BQ.1*, and XBB* Omicron subvariants (**Fig 1A, 1B**). We identified 434 infections involving multiple viral lineages and 85 additional cases consistent with reinfection within 90 days by a different lineage (see Methods) (**Fig S1**).

**Figure 1.**
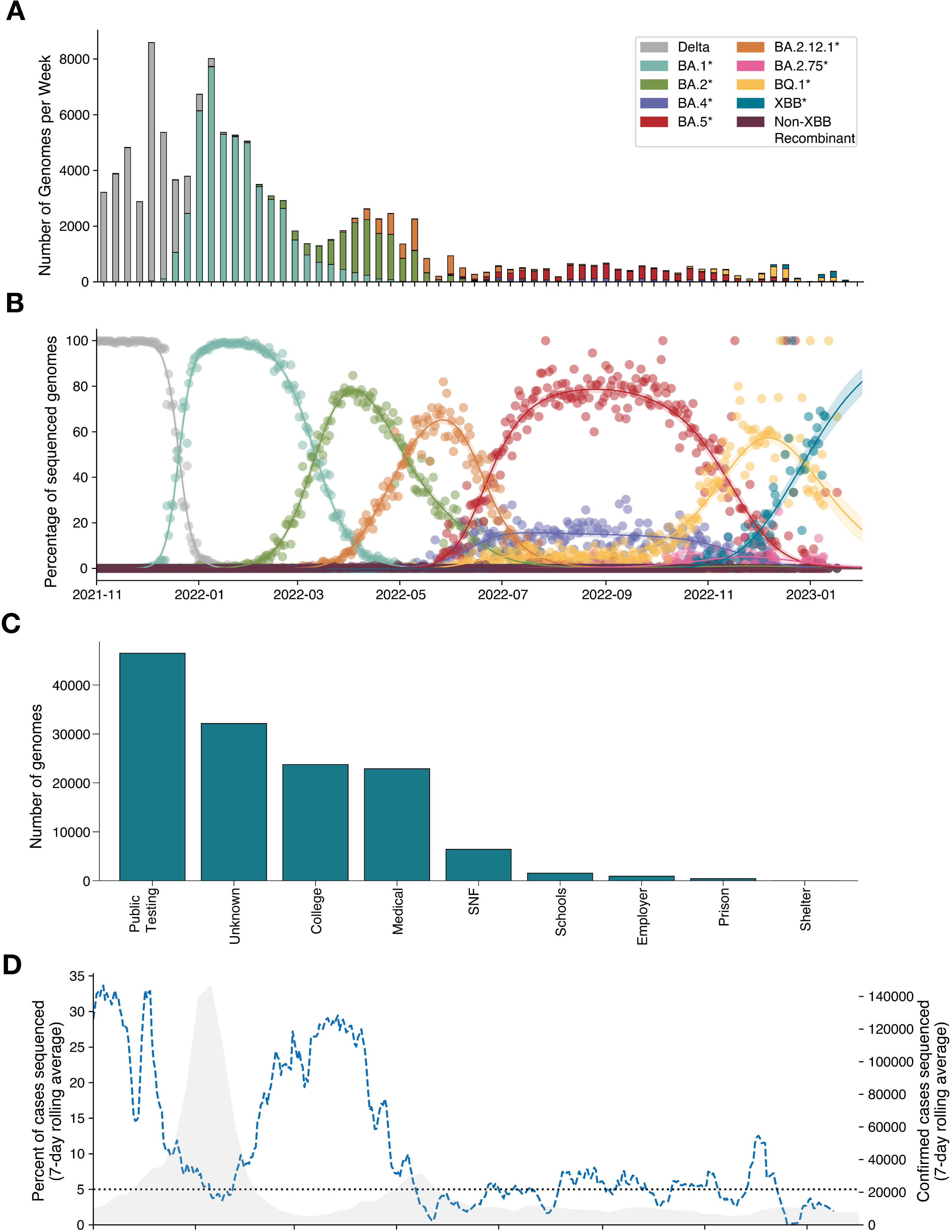
Study spans multiple viral lineages across diverse settings and demographic groups. A) Number of SARS-CoV-2 genomes sequenced per week attributed to each lineage in the complete dataset. The asterisk denotes that the lineage label (e.g., BA.1*) includes all sublineages. B) Multinomial regression modeling logistic growth of Delta and Omicron sublineages in the complete dataset. Shaded regions indicate 95% confidence intervals; points represent daily lineage frequencies from sequencing data. C) Number of sequenced genomes stratified by testing sector. D) Blue dashed line indicates the percentage of confirmed cases sequenced per day in this study shown as a 7-day rolling average (left y-axis). Gray shaded region shows SARS-CoV-2 case counts reported to Massachusetts Department of Public Health from November 1, 2021 to January 17, 2023 (7-day rolling average; right y-axis). The horizontal black line marks the United States Center for Disease Control and Prevention’s national target of sequencing ≥5% of confirmed cases.

The samples were collected at 666 facilities statewide, which we classified into seven sectors (**Fig 1C, Fig S2, S3**): 29 public testing facilities (accounting for 35% of genomes), 101 colleges (19%), 52 hospitals and clinics (17%), 301 skilled nursing, rehabilitation, and long-term care facilities collectively referred to as SNFs, (5%), 44 primary and secondary schools (1%), 23 other workplaces (<1%), and 117 testing facilities of unknown sector (22%). We obtained deidentified epidemiological data, including age, sex, city of residence, and vaccination information, for 85,125 total genomes from 81,952 unique individuals. Unless otherwise indicated, we used this set of genomes with accompanying epidemiological data as the core dataset for downstream analyses.

Sequenced samples broadly reflected the distribution of contributing testing facilities (**Fig S2-S4**) and tracked statewide case count trends, with a mean sampling fraction of 10.5% of all confirmed cases reported to MDPH per week during the study period (**Fig 1D**). Several sectors were enriched for specific demographic groups, including children in primary and secondary schools, young adults in colleges, and older adults in SNFs (**Fig S2D**). In contrast, free public testing was available statewide regardless of age or symptom status and therefore served as the sector most representative of the overall population (**Fig S4**).

### Distinct transmission patterns in different demographic groups

The diversity of environments and demographics represented by the testing sectors in our dataset allowed us to examine how these factors shaped SARS-CoV-2 transmission. We focused on settings with clear implications for shared exposure, including SNFs, colleges, primary and secondary schools, and a mixed group of other workplaces that included hospital staff, first responders, and warehouse staff. For each setting, we asked how much additional risk of infection was attributable to the shared facility environment compared to broader community risk.

We assessed within-facility transmission by quantifying the enrichment of closely related viral genomes within each setting compared to the surrounding community. That is, for each virus from a facility, we calculated how often other viruses from either the same facility or from the surrounding community were closely related to it, where the community was represented by public testing data from individuals in the same municipality. Increased within-facility viral relatedness therefore served as a proxy for the probability that transmission occurred within the facility rather than between unrelated individuals in the same municipality. For this purpose, closely related viruses were defined as those differing by ≤2 substitutions and collected within 10 days of one another, consistent with previous studies.^11–13^

Using this framework, we observed marked differences in within-facility transmission between settings. Closely related viruses were strongly enriched in SNFs (4.9x, 95% CI: 4.3–5.5x) and substantially enriched in colleges (3.3x, CI: 3.1–3.6x). In contrast, enrichment in schools was modest (1.4x, CI: 1.2–1.7x) and was not statistically significant in other workplaces (1.1x, CI: 0.9–1.3x) (**Fig 2A**). These patterns indicated that the contribution of within-facility transmission varied considerably by setting; notably, average viral relatedness in SNFs was nearly twice that observed in any other setting. These results were robust to differences in sampling density across sectors, as subsampling analyses yielded consistent enrichment patterns (**Fig S5**), and were not driven by a small number of highly sampled facilities (**Fig S6**).

**Figure 2.**
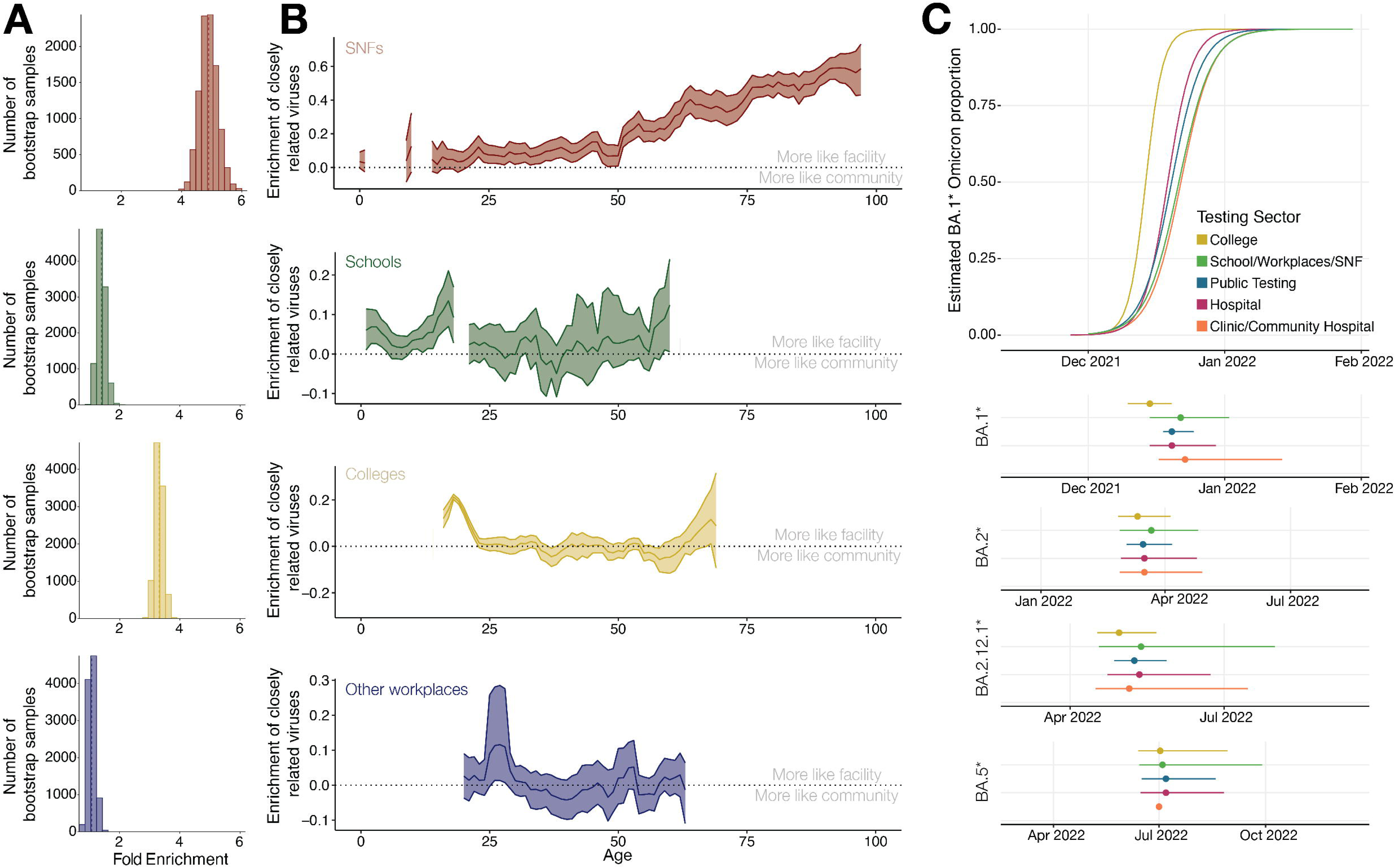
Heterogeneous transmission across facility types and age-defined subpopulations. A) Bootstrap distributions of fold-enrichment (x-axis) of closely related viruses within a facility relative to relatedness between the facility and surrounding municipality. The dashed line indicates the mean enrichment for each testing sector. B) Age-stratified enrichment of closely related viruses within each sector, defined as the difference in probability of a close viral relative within the same facility versus in public testing data from the individual’s municipality of residence. Shown for age groups with >30 samples (3-year rolling average; bootstrapped 95% confidence bands). C) Logistic regression of BA.1* rise over time (top) and estimated date at which lineage frequency reached 50% for BA.1*, BA.2*, BA.2.12.1*, and BA.5* across sectors (bottom, with 95% confidence intervals).

We also observed that in SNFs, schools, and colleges, enrichment of closely related viruses was largely confined to specific age groups. In SNFs, the probability that a virus had a close relative within the same facility increased steadily with age, beginning around 50 years old. Among the oldest individuals, viruses were 60 percentage points more likely to be closely related to another virus from the same facility than to one from the surrounding community (**Fig 2B**). Although our SNF dataset likely included some younger patients in rehabilitation and other care settings—so age does not perfectly distinguish staff from residents—the proportion of true residents increases with age. The strongest enrichment of closely related viruses therefore most likely reflects resident-to-resident transmission.

Examining within-facility and community relatedness separately provided additional insight into transmission in SNFs. Relatedness between viruses in SNFs and those in the community varied little by age, whereas relatedness among viruses in SNFs increased sharply with age (**Fig. S7**). This pattern is consistent with repeated viral introductions from the community into SNFs followed by relatively short transmission chains within facilities. Prolonged outbreaks from single introductions would instead be expected to generate longer transmission chains linking residents to community viruses, producing declining relatedness with age. These observations are not, however, informative about the frequency of introductions or overall incidence among staff and residents.

In schools, staff-aged individuals (defined here as >23 years old) showed the lowest within-facility enrichment of closely related viruses, lower even than in SNFs: their viruses were only 2% more likely to have a close relative in the school than in the broader community. Younger students (5-14 years old) also showed only modest enrichment (3%), with some variation by age. In contrast, enrichment increased sharply among older adolescents, peaking at ∼20% among 18-year-olds. Because both younger students and staff show little evidence of increased transmission, these findings suggest that the school environment itself conferred little additional risk. Instead, elevated transmission among older adolescents likely reflects age-specific social interaction patterns, whether inside or outside school.

Colleges showed a similar age-structured pattern, with enrichment of related viruses largely confined to undergraduates (typically 18 to 22 year olds). The strongest signal was among those 18–20 years old, for whom enrichment peaked at ∼20%. We found little evidence of increased viral relatedness among other age groups within colleges, including those typical of post-graduate students, faculty or staff. Together, the school and college analyses suggest that elevated SARS-CoV-2 transmission was not a general feature of these environments but was concentrated within a narrow age range. Consistent with this interpretation, public testing data also showed increased transmission among individuals in their late teens to early twenties (**Fig S8**), suggesting this age group contributed disproportionately to overall transmission, although the proportion who were college students is unknown.

The importance of young adults in colleges in SARS-CoV-2 transmission was further evident in examining lineage dynamics. We previously observed that BA.1 cases rose earlier and more rapidly in colleges than statewide^14^. Extending this analysis to major lineages for which we had sufficient data, BA.1*, BA.2*, BA.2.12.1* reached 50% frequency earlier in college settings than in other sectors, with lead times ranging from 4 to 13 days. BA.5* was an exception, it showed similar timing across all sectors, but rose when colleges were not in session (**Fig 2C**).

To disentangle effects of age from college settings, we compared 18- to 22-year-olds in college and public testing to all-age public testing for BA.1*, BA.2*, and BA.2.12.1* (omitting BA.5* for lack of data). BA.1* reached 50% frequency earliest among 18- to 22-year-olds in colleges, whereas BA.2* and BA.2.12.1* reached 50% frequency earlier among 18- to 22-year-olds in both colleges and public testing than in all-age public testing (**Fig S9**). These findings suggest that both the college environment and the 18–22 age group contributed to early lineage expansion, supporting the potential role of young adults as a sentinel population for genomic surveillance.

We considered the possibility that asymptomatic surveillance testing conducted in many colleges contributed to the initial rise of emerging variants there, since such testing could enrich for early detection of infections^14,15^. To test this, we grouped together the other sectors known to have been conducting predominantly asymptomatic surveillance testing (schools, workplaces, and SNFs) to increase statistical power. We then compared the rise of variants in this combined group with colleges, as well as with the remaining sector categories, (hospitals and clinics, and public testing), which performed mixtures of symptomatic and asymptomatic testing (**Fig S10**). As before, all variants except for BA.5* (which emerged when college was not in session) reached 50% frequency earliest in colleges. The consistent early rise of multiple lineages in colleges, coupled with the absence of this pattern in other sectors conducting asymptomatic surveillance, suggests that the early signal in college testing was not simply driven by asymptomatic testing. Instead, features of the college population and environment likely contributed to rapid early expansion, reinforcing the potential of colleges as settings for early viral detection.

### High resolution temporal and spatial mapping of viral spread

Our dataset captured the introduction of six major Omicron sub-lineages at high geographic resolution, enabling detailed analysis of how new variants were introduced, established, and spread. The dataset had broad coverage of the state, including genomes from ≥1% of all confirmed cases in 87% (304 of 351) of municipalities (**Fig 3A**). To quantify viral introductions and movement, we reconstructed phylogenetic trees using two complementary sampling strategies: one normalized to case counts and one based on equal sample sizes per week per municipality. We defined an introduction into the state as a virus detected in our dataset whose immediate ancestral node was inferred to be outside Massachusetts. Similarly, we identified intrastate movement when the inferred locations of ancestral and descendant nodes shifted between municipalities within the state.

**Figure 3.**
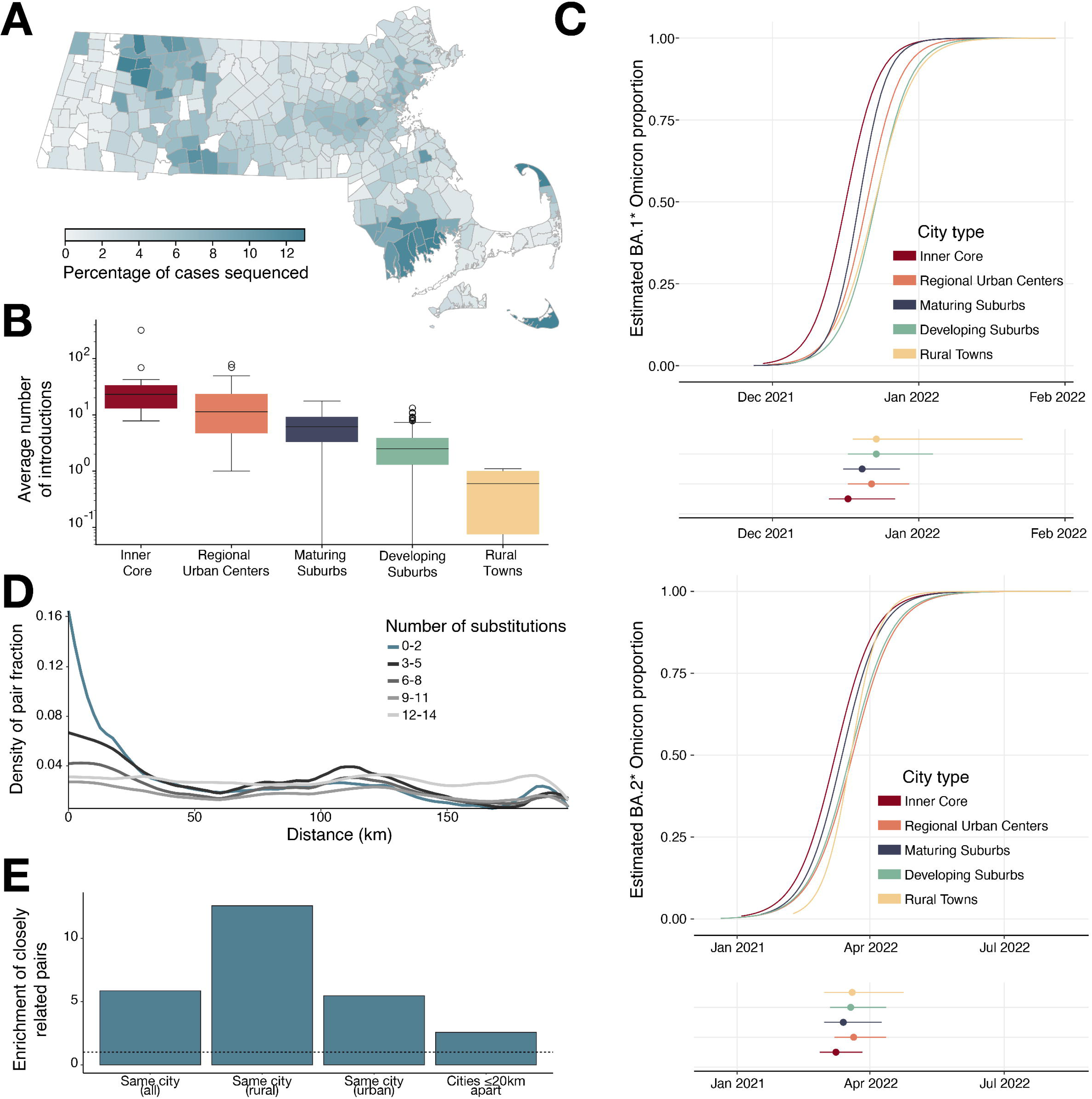
Viral movement within Massachusetts driven by larger and denser populations. A) Percentage of confirmed cases sequenced in each municipality during the study period. B) Mean number of inferred introductions into each municipality, averaged across 10 phylogenetic trees and all six major lineages, stratified by municipality type. C) Logistic regression of BA.1* (top) and BA.2* (bottom) lineage frequency over time, showing estimated date at which lineage reached 50% frequency in municipalities of different urbanicity classes (95% confidence intervals). D) Proportion of virus pairs within specified genetic distance (as a fraction of all pairs) across geographic distance between municipalities of residence. Closely related pairs (≤2 substitutions) are shown in blue. E) Relative enrichment of closely related virus pairs across different municipal settings. “Rural” includes rural municipalities and developing suburbs; “urban” includes maturing suburbs, regional urban centers, and the inner urban core. The dashed line indicates background relatedness for each category.

New introductions into the state were disproportionately likely to occur into more urban areas. That is, even when municipalities were equally sampled, the rate of introductions was significantly and independently associated with both the size and the density of the municipality’s population, suggesting greater per-capita linkage to out-of-state travel in urban areas (**Fig S11**). Movement of viruses between municipalities was similarly associated with both the size and density of the source population. As a result, urban municipalities acted as both recipients of new introductions from outside the state and sources of intrastate spread (**Fig 3B**). Consistent with this, we found that BA.1* and BA.2* (the only lineages with enough data) rose to high frequency initially in the primary urban center, followed by the surrounding suburbs and then by smaller independent urban areas, with more rural areas trailing (**Fig 3C**).

We estimated the speed of viral movement throughout the state by looking at how genetic distance varied at different geographic distances. For pairs of viruses collected within 10 days of each other, the number of genetic differences between them should be proportional, on average, to the number of intervening transmissions since the pair’s common ancestor. Closely related viruses (≤2 substitutions) occurred predominantly at short physical distances (<∼15 km) (**Fig 3D**). By the time a pair of viruses were separated by 3-5 substitutions (corresponding to ∼4 generations or 20 days of transmission since their common ancestor), they were almost as likely to be found >100 km apart as within the same municipality. A substantial component of this spread involved movement between major urban centers. For example, the relatedness peak in **Fig 3D** around 125 km, which is most pronounced for a genetic distance of 3–5 substitutions, reflects the distance between two major, well-sampled cities (Boston and Springfield); similar patterns were observed for other metropolitan pairs. In contrast, pairs involving at least one virus from a more rural municipality consistently showed lower relatedness across distances, consistent with viruses primarily moving between urban centers rather than from urban to rural municipalities. When genetic distance exceeded ∼12 substitutions, geographic structure became undetectable, suggesting that descendant viruses typically diffused broadly throughout the state within roughly two months.

Although viruses in rural areas were less likely to be closely related to those from distant municipalities, they were more likely to be closely related to other viruses from the same community. In developing suburbs and rural municipalities, viruses from the same municipality were 11.3x as likely to be closely related as viruses from different municipalities, compared to only 5.6x in the inner core, regional urban centers, and maturing suburbs (**Fig 3E**). Greater relatedness in less populous towns may reflect both the greater isolation from intrastate movement and smaller effective viral population sizes in those settings. These different aspects of the genomic data, then, create a consistent picture, in which new variants typically arrived first in urban areas, spread rapidly to adjacent suburbs and then to other regional urban centers, and reached rural areas more slowly and less completely.

### Vaccination was associated with decreased infection and transmission

Although this opportunistically collected dataset was not designed to directly assess the effects of vaccination on infection or transmission, we leveraged the available data to explore these relationships and observed modest but informative associations. In one cohort, the timing of the COVID-19 vaccine rollout allowed us to examine the qualitative impact of vaccination on infection. Children aged 5–11 years old first became eligible for vaccination in the US on October 29, 2021, two days before the start of our study, with administration of first doses ramping up in mid-November (**Fig S12A**) and second doses following in mid-December. Comparing the periods immediately before and after widespread vaccination (November vs. late December/early January), we found that the proportion of infected individuals in the 5- to11-year-old cohort declined by approximately 50% in both school-based and public testing data **(Fig. 4A,B)**, with the decline evident for both Delta and BA.1* **(Fig. S12B)**. The concordance between school-based and public testing suggests that vaccination provided substantial short-term protection against infection.This protection may have contributed to the relatively low within-school transmission among younger students noted above, although it is unlikely to be the dominant factor, since similarly low values were observed among 12- to 14-year-olds.

**Figure 4.**
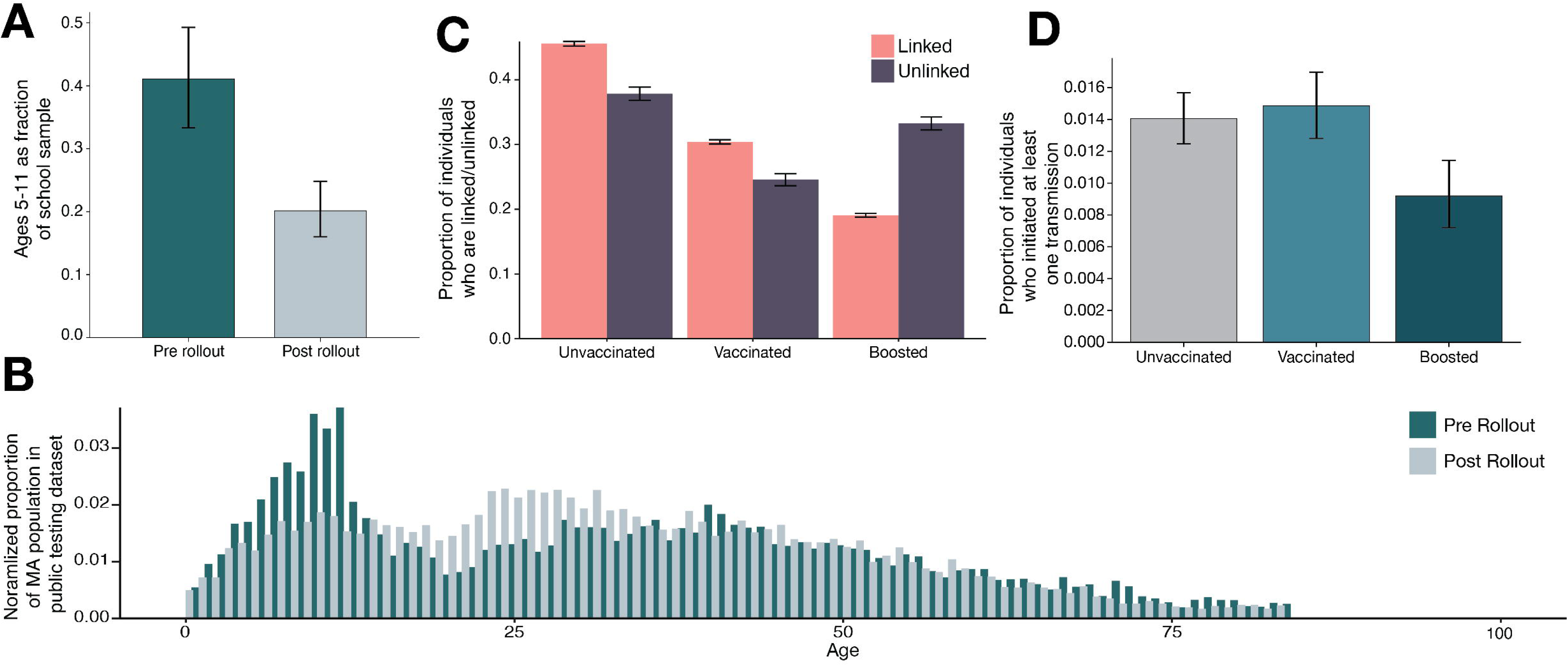
Booster vaccination is associated with reduced onward SARS-CoV-2 transmission. A) Proportion of individuals in school-based testing who were 5–11 years old, comparing November 2021 with late December 2021/early January 2022. B) Fraction of the Massachusetts population represented in the public testing dataset by age during November 2021 (blue) versus late December 2021/early January 2022 (gray). Plots are normalized to have equal areas. C) Proportion of linked (coral) and unlinked (dark purple) cases across vaccination categories in the complete dataset; linked individuals have closely related viral genomes elsewhere in the dataset and are therefore more likely to have been involved in transmission chains. D) Proportion of individuals with a detected descendant virus as, identified through shared intrahost single nucleotide variants (iSNVs), stratified by vaccination status in the public testing dataset.

We asked whether vaccination altered SARS-CoV-2 transmission between individuals using two complementary genomic approaches. Both analyses suggested an association between booster vaccination and reduced onward spread. The first approach distinguished likely transmitters from non-transmitters by proxy, classifying individuals whose viruses had closely related genomes elsewhere in the dataset as “linked”, i.e. as evidence of a short transmission chain between them, and those without such relatives as “unlinked”. Unlinked individuals were significantly more likely to have received a booster than linked individuals (33.2% vs. 19.1%; **Fig 4C**), indicating a weaker connection between boosted individuals and sampled transmission chains. The second approach identified likely transmitters by detecting within-host variants that later appeared as consensus variants in other individuals, consistent with onward transmission^16^. In this analysis, boosted individuals were 0.65x (0.50–0.84) as likely as unvaccinated individuals to initiate at least one transmission event (**Fig 4D**). In contrast, we observed no significant differences between unvaccinated individuals and those who had completed a primary vaccine series without a booster in either analysis.

We next asked whether the association between booster vaccination and reduced onward spread reflected recency of vaccination rather than number of vaccine doses received. To test this, we compared unboosted individuals who were first vaccinated early in the initial rollout (February-May 2021) with those first vaccinated later (November 2021- January 2023).

Individuals vaccinated later showed a 0.85x (0.54x–1.33x) likelihood of initiating transmission relative to unvaccinated individuals (**Fig S13**). Although this difference was not statistically significant, the trend is consistent with previous observations of waning vaccination protection against SARS-CoV-2 transmission^17^. We also observed lower viral loads (based on diagnostic Ct values) with increasing number of vaccine doses, and a modest but statistically significant increase in viral load with time since the most recent vaccine dose^18^ (**Fig S14**). Together, these results suggest that the increased probability of transmission with time since vaccination may be partly mediated by biological factors related to viral load. However, both the number and timing of vaccine doses are also likely correlated with behavioral factors that influence transmission probability, such as risk tolerance or the ability to self-isolate.

### Sampling rates for detection of novel lineages

During the pandemic, practical questions arose about the rate of viral sequencing needed to effectively surveil emerging variants of concern. Using our dataset of all 130,000 genomes, we first asked how sampling rate influences the speed of detecting new variants. Using both a simple binomial model and the tool PhyloSamp^19^, we modeled the detection of lineages with the observed growth rates of BA.1*, BA.2*, BA.2.12.1*, BA.5*, and BQ.1* (the lineages with sufficient data). Specifically, we assessed how the weekly sampling rates affected the timing of first detection relative to the date each variant reached 1% prevalence in the dataset, a commonly used surveillance threshold^20,21^. Sequencing 100 genomes per week resulted in detection before the 1% threshold more than 50% of the time, while 50 genomes/week typically delayed detection by only a few days. The probability of detection increased with increasing sequencing rate up to approximately 500 genomes per week. At this rate, four of the five lineages (BA.2*, BA.2.12.1*, BA.5*, and BQ.1*) had >95% probability of being detected by the target date, with little additional benefit from higher sampling (**Fig 5A, Fig S15**). In contrast, BA.1* rose so rapidly in frequency that sampling intensity had little effect on detection timing; increased sampling would not have enabled earlier detection.

**Figure 5.**
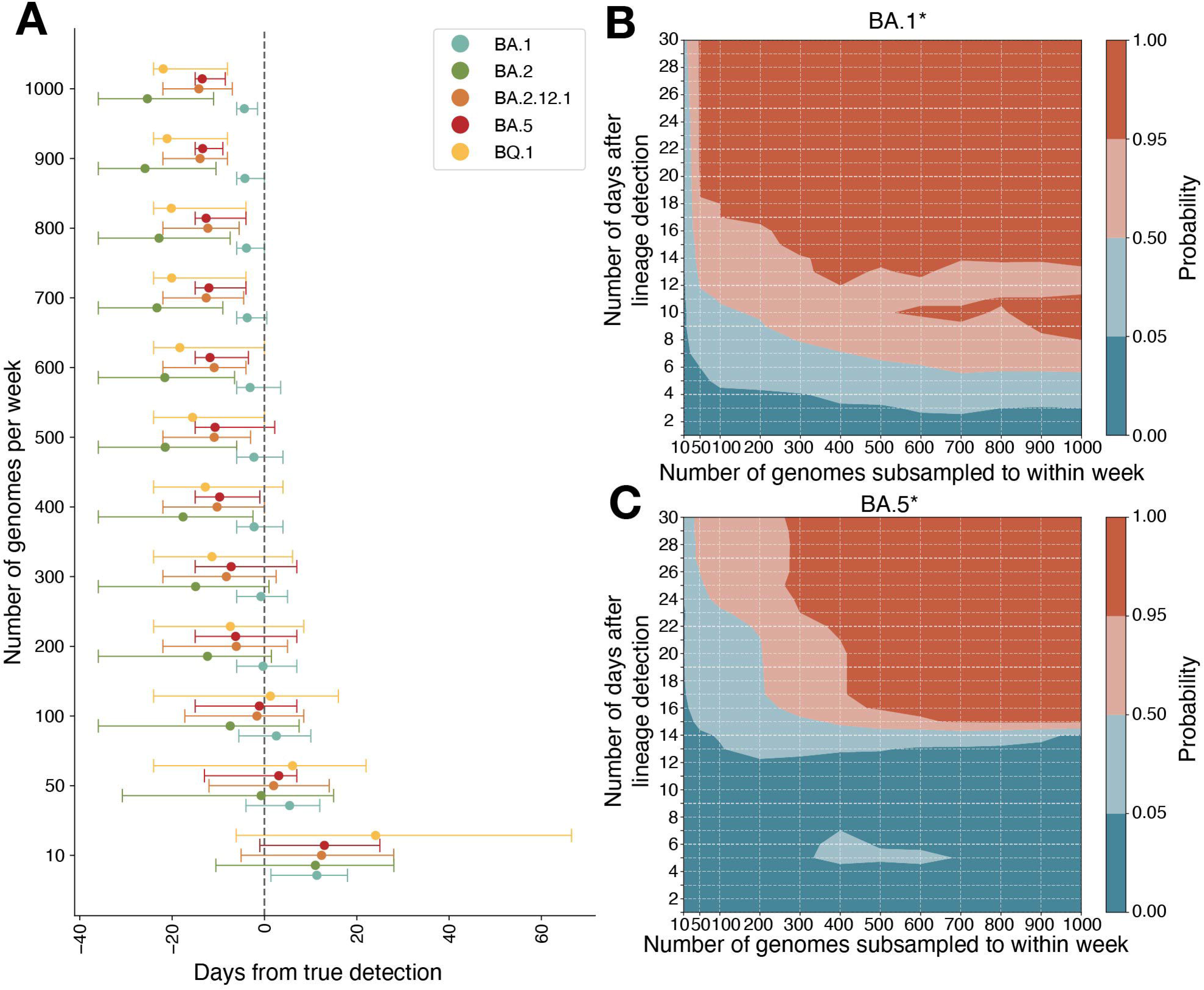
Genomic sampling intensity determines the speed of variant detection. A) Estimated timing of detection of a rising variant relative to the date it reaches 1% frequency in the dataset, across different weekly sampling rates (95% credible intervals). B) Subsampling analysis of BA.1* empirical data at N genomes per week. Logistic regression models were fit beginning one day after BA.1* reached 3% frequency in the dataset. Shown is the probability that the 95% confidence interval of the estimated growth rate exceeds 0, indicating strong evidence for logistic growth. Red colors denote probabilities >50%. C) Same as in (B), applied to BA.5*.

We next estimated the sampling rate required for a more data-intensive surveillance goal: identifying previously unknown lineages that are increasing in frequency and therefore represent potential variants of concern. To do this, we repeatedly subsampled our dataset of 130,000 genomes for the same five lineages at increasing weekly sampling rates. For each subsample, we fit logistic regression models beginning on the day the lineage reached 3% frequency and extending over varying time windows to determine the earliest date at which the growth rate became significantly positive (95% confidence). The 3% threshold minimizes the highly stochastic fluctuations that occur prior to the onset of exponential growth. Based on these empirical data, sequencing 300 genomes per week was sufficient to determine within 14 days that BA.1* was increasing in frequency **(Fig. 5B)**. In contrast, for slower-growing variants such as BA.5*, confident detection of growth within 14 days was effectively unattainable; even detection within 21 days would have required sequencing at least 400 genomes per week **(Fig. 5C)**.

## DISCUSSION

Large-scale genomic surveillance provided valuable insights into the emergence and spread of SARS-CoV-2 variants during the COVID-19 pandemic, but deeper epidemiological analyses were often hindered by the lack of detailed metadata linked to viral genomes. Here, we leverage large-scale, high-resolution SARS-CoV-2 genomic surveillance data paired with epidemiological metadata across multiple variant waves to examine spatial patterns of viral spread, differences in transmission dynamics across demographic groups and settings, and factors associated with the risk of infection and onward transmission. We also provide practical estimates of the sampling rate required for timely detection of known variants of concern and for identifying previously unknown variants that are increasing in frequency.

Within schools, colleges, and SNFs, elevated SARS-CoV-2 transmission relative to public testing was concentrated in specific age-defined subpopulations rather than reflecting a general feature of these institutional settings. Notably, in all three settings, transmission involving staff-aged individuals was only modestly higher than, or indistinguishable from, transmission in the surrounding community. We also found no evidence of substantially elevated transmission in a heterogeneous group of other workplaces, nor among children under the age of 15 in schools. Although sample sizes for schools were relatively small in our dataset, the essential findings, particularly the importance of young adults and the lower transmission risk among staff, were consistent across sectors. Together, these results suggest that increased infection risk in these settings was largely driven by characteristics of specific subpopulations rather than by the institutional settings themselves.

The demographic groups that showed elevated transmission likely reflect different processes in their respective contexts. SNFs were the only setting that showed a consistent enrichment of within-facility transmission. The substantial increase among older age groups suggests a unique risk to residents in these facilities, while the more modest increase among individuals under 50 could indicate increased risk among staff, although it may also reflect younger in-patients in rehabilitation or other care settings.

In schools, the sharp increase in transmission was largely confined to 15- to 18-year-olds, suggesting that it reflects distinctive social interaction patterns in this age group, only some of which occur within the school environment. It was somewhat surprising that we did not observe stronger evidence for school-associated transmission across a broader age range, given the well-established role of younger children in the spread of endemic respiratory viruses, the many opportunities for transmission within schools and school-adjacent environments, and the additional shared risk associated with siblings attending the same school. A particularly notable and reassuring finding was how limited the apparent workplace risk for teachers was, at least under the conditions present in our data.

We observed similar age-structured transmission among the undergraduate-aged population in college settings. The particularly sharp increase between ages 18 and 20 may reflect the additional effect of shared living environments, as on campus college dormitories are more common in this age group than among older students who typically live off campus. Whatever the underlying causes, evidence for elevated transmission among young adults comes from both college and school populations, groups that typically have limited overlap in their social networks.

Our findings here and elsewhere^14^ suggest that individuals aged 18 to 22 served as leading indicators for the establishment of new SARS-CoV-2 lineages; emerging variants expanded within young adult populations before becoming widespread across the rest of the state, with the effect strongest among college students. Because Massachusetts has a high concentration of colleges and other post secondary institutions, we considered whether an overrepresentation of young adults, particularly residential college students, might influence this pattern. We were unable to readily identify data directly comparing the proportion of residential college students in Massachusetts with that of the U.S. overall. However, U.S. Census Bureau data indicate that 18- to 22-year-olds constitute a very similar share of the total population in Massachusetts and nationwide, approximately 6.5% in both cases^22,23^. (We note that the Census counts college students where they live during the academic year, whether on or off campus, even if they are originally from another state). In any case, our observation reflects the relative transmission rate within this demographic group rather than its share of the population, a relationship likely to hold in other contexts.

Although a genomic epidemiology dataset of this scale and resolution would ideally enable more extensive evaluation of public health interventions on transmission and infection risk, the timing and nature of interventions during our study period largely precluded such analysis. By the start of the study, many earlier widespread measures, including masking and distancing mandates and broad school closures, had already ended, leaving vaccination as the primary public health intervention for most of the period. Early in the study, some masking and other mitigation measures were intermittently in place, but these coincided with rapid shifts in the immune landscape as Delta was replaced by the BA.1 Omicron surge, along with extensive travel and holiday-related school and college closures. As a result, we were unable to reliably isolate the effects of specific policy changes on transmission.

Nevertheless, we observed a short-term reduction in infections among 5- to 11-year-olds coinciding with vaccine rollout, detectable in both school-based surveillance and broader public testing. We were unable to estimate the effect of vaccination on infection more broadly because we did not have vaccination data for uninfected people to use as a comparator. Assessing vaccination effects on transmission was more tractable, as these could be inferred from viral genetic relationships combined with vaccination status metadata within our dataset. Individuals with more recent and additional vaccine doses had a lower probability of initiating transmission, although causal interpretation remains limited by potential confounding factors, such as behavioral differences between groups.

The arrival of multiple new lineages during our study period, each effectively representing a new outbreak, allowed us to examine patterns in where lineages were introduced, how they subsequently spread, and how they behaved across different populations and settings. New lineages were introduced primarily through major urban areas, from which they spread quickly to nearby suburbs and regional centers, and more gradually to less populated areas, becoming broadly distributed statewide after approximately two months. These patterns suggest that effective strategies to reduce transmission may differ by geographic density, for example, prioritizing efforts to limit new introductions into urban centers while focusing on interrupting local transmission chains in rural areas.

Our analyses also clarify how sequencing coverage influences the timeliness of variant detection. Increasing the fraction of cases sequenced reduces the delay to detecting emerging lineages, but with diminishing returns at higher coverage levels. These results highlight a practical tradeoff between surveillance effort and detection speed, suggesting that sustained sequencing at moderate coverage can capture much of the achievable early-warning benefit.

Despite the constraints inherent to opportunistic genomic surveillance data collected during a rapidly evolving pandemic, this study illustrates the power of integrating genomic and epidemiological information to resolve fine-scale transmission patterns. By identifying which groups were most likely to drive transmission within and across settings, and when emerging variants were most likely to become detectable, our findings provide actionable insights for the design of future genomic surveillance systems, particularly through targeted sampling of high-transmission demographic groups.

### Limitations of study

While offering important novel findings, this study was subject to common limitations of genomic epidemiology, including opportunistic, observational datasets, siloed data streams, and the broader constraints of a rapidly evolving public health emergency. In particular, our sampling framework was shaped by the realities of a large-scale COVID-19 testing program and was not designed to resolve details of transmission dynamics. Although epidemiological metadata, rarely available at this scale, enabled comparisons across demographic groups and settings, it did not permit definitive linkage of infections to individual behaviors or specific exposure events, limiting causal inference. These constraints also restricted our ability to distinguish the behavioral and biological drivers underlying the observed patterns. For example, exposure risk and testing practices may vary in correlated ways between facility types, and vaccination status may itself be associated with differences in risk-taking behavior, not captured in this study. In addition, clinical data were available for only a subset of individuals and are often complex and subjective, limiting the scope of related analyses, while vaccination status was unavailable for uninfected individuals, precluding more detailed assessment of vaccine-associated effects. As a result, analyses requiring deeper integration of clinical or behavioral data could not be pursued robustly.

## Data Availability

All data produced in the present study are available upon reasonable request to the authors

## Acknowledgements

We thank members of the Massachusetts Department of Public Health and Broad Clinical Labs (formerly Broad’s Clinical Research Sequencing Platform) for their efforts and support of this work. We are also grateful to the laboratories that shared SARS-CoV-2 sequence data publicly in GenBank. We thank Miguel Paredes for helpful feedback on the draft manuscript and analyses.

## Funding

Samples analyzed were from CDC-funded COVID-19 diagnostic testing and genomic surveillance programs (75D30121C10501 and 75D30122C13720 to the Broad Institute Clinical Research Sequencing Platform, LLC) and through the U.S. CDC Pathogen Genomic Centers of Excellence (INTF5104H78W22195346 to P.C.S. and B.L.M.). This work was also funded by a CDC Broad Agency Announcement (75D30120C09605 to B.L.M.), a Howard Hughes Medical Institute Investigator Award to P.C.S., the National Institute of Allergy and Infectious Diseases (U19AI110818, to P.C.S.), the Rockefeller Foundation (2021HTH013 to B.L.M. and P.C.S.), the National Human Genome Research Institute (T32HG010464-06 to G.K.M.), the National Institute of General Medical Sciences (T32GM007753 and T32GM144273 to B.P.), and the Broad MD-PhD Fellowship (to B.P.).

## Competing interests

P.C.S. is a co-founder of, shareholder in Delve Bio; she was formerly a co-founder of and shareholder in Sherlock Biosciences, Inc and a Board member of and shareholder in Danaher Corporation.

## Author contributions

Conceptualization: G.K.M., T.B.-F., L.A.K., S.F.S., C.M.B., B.L.M., P.C.S.

Methodology: G.K.M., T.B.-F., L.A.K., S.F.S., M.B., B.P., I.S., S.W., D.J.P.

Software: G.K.M., T.B.-F., L.A.K., S.F.S., M.B., C.E.C., K.S.M., B.P., I.S., S.W., D.J.P.

Formal analysis: G.K.M., T.B.-F., L.A.K., S.F.S., M.B., C.E.C., K.S.M., B.P., I.S., S.W., D.J.P.

Investigation: G.K.M., T.B.-F., L.A.K., M.B., C.E.C., K.S.M., B.P., I.S., K.C.D., K.J.S., C.L., N.A.F., H.M.R.

Resources: S.B.G., S.S., S.W., L.C.M., C.M.B., B.L.M., P.C.S.

Data curation: G.K.M., T.B.-F., L.A.K., M.B., C.E.C., K.S.M., N.A.F., H.M.R.

Writing – original draft: G.K.M., T.B.-F., L.A.K., S.F.S., B.L.M.

Writing – review & editing: G.K.M., T.B.-F., L.A.K., S.F.S., B.L.M., P.C.S.

Supervision: G.K.M., S.F.S., D.J.P., L.C.M., C.M.B., B.L.M., P.C.S.

Project administration: G.K.M., T.B.-F., L.A.K., C.M.B., B.L.M.

Funding acquisition: C.M.B., L.C.M., P.C.S., B.L.M.

All authors reviewed and approved the manuscript.

## Declaration of interests

B.L.M. and P.C.S hold patents related to genomic sequencing technologies. P.C.S. is a co-founder of, shareholder in Delve Bio and Lyra Labs; she was formerly a co-founder of and shareholder in Sherlock Biosciences, Inc and a Board member of and shareholder in Danaher Corporation.

## Declaration of generative AI and AI-assisted technologies in the writing process

During the preparation of this work the author(s) used ChatGPT in order to refine language in the writing process. After using this tool/service, the author(s) reviewed and edited the content as needed and take(s) full responsibility for the content of the published article.

## STAR Methods

### RESOURCE AVAILABILITY

#### Lead Contact

Further information and requests for resources and reagents should be directed to and will be fulfilled by the lead contact, Gage K. Moreno.

## Materials availability

This study did not generate new unique reagents.

## Data and code availability

- Raw reads for all samples (including those that did not produce a successful genome) were deposited in NCBI SRA. All NCBI data were deposited under BioProject PRJNA715749.
- All code used for sequence data processing, genome assembly, and phylogenetic analysis is publicly available either via the Dockstore Tool Registry Service (<u>dockstore.org/organizations/BroadInstitute/collections/pgs</u>) or on GitHub (<u>github.com/AndrewLangvt/genomic_analyses/blob/main/workflows/wf_viral_refbased_assembly.wdl</u>).
- For any additional information required to reanalyze the data reported in this paper, contact the lead author.

## EXPERIMENTAL MODEL AND SUBJECT DETAILS

### Ethical approvals

As previously described^5^, the research project (Protocol #1603078) was reviewed and approved by the Massachusetts Department of Public Health (MADPH) Institutional Review Board and covered by a reliance agreement at the Broad Institute. An additional non-human subjects research determination was made by the Harvard Longwood Campus Institutional Review Board. A summary of all available metadata categories and the number of individuals within each category is provided in **Table 1**.

## METHOD DETAILS

### SARS-CoV-2 RT-qPCR

For specimens submitted for testing under Emergency Use Authorization to the Broad Institute’s CLIA-certified laboratory, Clinical Research Sequencing Platform (now Broad Clinical Labs), total RNA was extracted from inactivated anterior nasal (AN) swabs using the Thermo Fisher MagMAX Viral RNA Isolation kit and presence of virus was confirmed by RT-qPCR assay detecting the N1 and N2 SARS-CoV-2 gene regions. Ct values for the N1 gene were used to compare viral titers between individuals; samples for which the RP positive control gene had a Ct>32 were excluded from the analyses to prevent biasing from poorly collected samples. A specimen was determined positive for SARS-CoV-2 if the initial cycle threshold of the N1 probe was less than or equal to Ct 40. As seen in previous studies^24,25^, clinical diagnostic PCR cycle threshold (Ct) values were significantly lower for Delta than for the Omicron sublineages, with little variation among the latter (**Fig S14**).

### SARS-CoV-2 sequencing

Candidate samples were re-extracted from the source material for Illumina sequencing. Libraries were prepared using the NEBNext ARTIC SARS-CoV-2 FS Library Prep Kit and transitioned through ARTIC, VarSkip Short v1, v1a, or v2a amplicon primers to ensure genomic coverage of emerging variants. During library preparation, some volumes were adjusted from manufacturer recommendations to accommodate 384-well plate reactions and high-throughput automated processing. Libraries were sequenced on Novaseq SP flowcells with 75-nucleotide paired-end reads.

### Genome assembly and analysis

For genomes generated at the Broad Institute, we conducted all analyses using viral-ngs 2.1.28 on the Terra platform (app.terra.bio). All of the workflows named below are publicly available via the Dockstore Tool Registry Service (dockstore.org/organizations/BroadInstitute/collections/pgs). Briefly, samples were demultiplexed, reads were filtered for known sequencing contaminants, and SARS-CoV-2 reads were assembled using a reference-based assembly approach with the SARS-CoV-2 isolate Wuhan-Hu-1 reference genome NC_045512.2 (using workflow sarscov2_illumina_full.wdl; publicly available on GitHub). We processed all raw read data using a reference-based consensus calling method with the same NC_045512.2 reference genome. Assembled genomes meeting the CDC criteria for submission to public repositories (unambiguous length ≥24,000 nt and successful gene annotation) were deposited in NCBI Genbank and GISAID immediately upon completion. Raw reads for all samples (including those that did not produce a successful genome) were deposited in NCBI SRA. All NCBI data were deposited under BioProject PRJNA715749.

We used LoFreq version 2.1.5 to call intrahost single nucleotide variants (iSNVs) with default parameters (minimum read depth ≥10, strand bias <85%, and default iSNV quality scoring)^26^.

Variants with frequency ≥3% and in positions with read depth ≥100 were used for downstream analysis. Minor variants arising in primer binding sites were masked^27^. All non-fixed consensus mutations (<97% and >50%) were flipped to their corresponding minor variant (e.g. a mutation at C11803T at 65%, would now show up as a T11803C at 35%). An additional filtering step was applied to account for sequencing bias^28,29^. Briefly, a binomial distribution was applied that took into account the total number of variant reads, sequencing depth, and sequencing error rate.

Variants with significance (≤0.05) after applying a Bernoulli correction were kept for downstream analysis^29^.

### Identification of reinfections and coinfections

Among the 81,952 unique individuals, 2,714 had more than one genome sequence available within a 90-day window, corresponding to the timeframe during which repeated tests per individual could be tracked in the Massachusetts public health database (**Fig S1**). For individuals with multiple sequenced samples, we recorded the pairwise genetic distances between consensus genomes and the number of days between sample collections. Each genome was assigned a Pango lineage using pangolin (v4.1.3). Samples collected more than 14 days apart that were assigned to different Pango lineages were classified as reinfections, according to the CDC definition^30^. Individuals with multiple samples collected within 14 days were compared for lineage consistency; those showing identical lineage assignments were considered to have the same infection, while those with discordant lineages were classified as potential multiple infections within a short interval. Pairwise distances and collection intervals were visualized and colored by Pango lineage to confirm lineage differences and reinfection events (**Fig S1**).

To detect simultaneous infection with multiple viral lineages, we first assigned a consensus Pango lineage to each genome using pangolin (v4.1.3). We then compiled a list of major lineage-defining mutations for all major lineages (BA.1, BA.2, BA.2.12.1, BA.5, BQ.1, XBB). Samples were classified as co-infections if two or more lineage-defining mutations from a different lineage were present as iSNVs. Only samples with sufficient genome coverage (>90% completeness) and sequencing depth (>100x mean coverage) were considered to minimize false positives. The majority of identified co-infections involved Delta and BA.1 lineages, consistent with the high transmission period during the early BA.1 wave.

### Phylogenetic tree construction

All genomes paired with metadata during the study period (November 1, 2021, to January 17, 2023) were used for phylogenetic inference (n=94,404). Prior to phylogenetic inference, genomes were aligned using Nextclade CLI v1.3.0. Genomes were excluded if they 1) contained any mixed bases, 2) were missing >10% of the SARS-CoV-2 genome, 3) lacked adequate metadata, or 4) were denoted as “bad” during Nextclade QC. The remaining (n=91,080) genomes were masked using a light masking scheme^31^.

To understand the number of introductions into each municipality, we generated 60 variant-specific (10 per variant) maximum likelihood phylogenetic trees using publicly available SARS-CoV-2 genomes from Genbank (retrieved January 23, 2024). Datasets were filtered to only include genomes collected prior to the date that each major lineage reached 50% frequency in MA. The resulting dataset was further subsampled so that 1% of confirmed infections were sequenced within each geographic resolution (country, state, municipality), as the geographic distribution of cases in Massachusetts was skewed towards highly urban areas. We constructed a maximum-likelihood (ML) phylogenetic tree using FastTree^32^ with 100 bootstraps and otherwise default parameters.

Under this case-normalized sampling strategy, replicate tree sizes varied modestly by lineage: BA.1 trees contained 72,356–72,746 genomes; BA.2.12.1 trees contained 8,721–8,803 genomes; BA.5 trees contained 39,368–39,717 genomes; BQ.1 trees contained 6,605–6,772 genomes; and XBB trees contained 1,893–1,933 genomes.

We generated a set of temporally balanced subsampled phylogenetic trees. First, using the augur filter tool, we randomly sampled 2,000 genomes per month from the USA and from the global dataset, respectively, prioritizing sequences with close genetic similarity to samples in our study. Second, we included 5 genomes per week from each Massachusetts municipality with an average sequencing rate of at least five genomes per week.

Under this temporally balanced strategy, final tree sizes were smaller and more uniform across lineages: BA.1 trees contained 9,601 genomes (5,840 from Massachusetts); BA.2 trees contained 4,211 genomes (1,129 from Massachusetts); BA.2.12.1 trees contained 4,206 genomes (468 from Massachusetts); BA.5 trees contained 3,616 genomes (542 from Massachusetts); BQ.1 trees contained 3,609 genomes (147 from Massachusetts); and XBB trees contained 3,985 genomes (75 from Massachusetts).

Using custom scripts, the resulting phylogenies were processed using the refine, ancestral, traits, and export steps of the Nextstrain pipeline^33^.

### Viral introductions

To identify viral introductions into Massachusetts municipalities, we assigned a trait to each genome in the phylogeny of either “out-of-state” or the name of the municipality. We used Nextstrain’s ancestral inference^33^ to infer the state of that trait for each internal node of the tree.

We defined an introduction by traversing the phylogenetic tree using baltic^34^, identifying branches in which an ancestral node was inferred to be out of state with ≥70% confidence and gave rise to a descendant node inferred to be a Massachusetts municipality with ≥70% confidence.

For viral movements occurring between Massachusetts municipalities, we applied the same logic, defining a viral movement between municipalities as instances of an ancestral node and descendant node being different municipalities, both with confidence ≥70%. We combined the movement data between origin and destination municipalities with associated variables such as case counts, percent positivity, and population size and density for both origin and destination. Movements across the 10 replicate phylogenies for each variant were deduplicated on the origin, destination, date, and downstream genomes. The remaining movements were then counted based on origin, destination, and date. We used a Generalized Linear Model (GLM) with a Gaussian family and identity link function to examine the relationship between movement counts and various predictors. The model was specified as:

Movement_count LL0+L1(predictors) + L

Model fitting was performed using the Iteratively Reweighted Least Squares (IRLS) method in the statsmodels library v0.14.2 ^35^ in python. Model performance was assessed using the Akaike Information Criterion (AIC) and log-likelihood.

### Normalized age distribution pre- and post-vaccine rollout, ages 5–11

The age distribution was calculated for all public testing samples with submittable genomes and recorded subject ages during two periods, November 1, 2021 – November 30, 2021 and December 15, 2021 – January 13, 2022. The statewide population by age was interpolated using a cubic spline from US Census estimated data for 2021 ( https://www.census.gov/data/tables/time-series/demo/popest/2020s-state-detail.html). The two distributions are normalized to have equal areas.

### Pairwise analysis

First, all possible pairs of genomes within the same lineage, collected within 10 days of each other in the public testing sector were identified. Then, the genetic distance between all possible pairs, defined as the Hamming distance between genomes only including positions at which both genomes had confident reads, was calculated using the ape^36^ package v5.7.1 in R. Additionally, we similarly calculated all pairs among all genomes collected at SNFs, other workplaces, schools, and colleges.

### Geographic analysis

For all resulting genome pairs, we calculated geographic distance (calculated as haversine distance) between the interior points of each pair of municipalities with the R package *geosphere*^37^ V 1.5.18. Geographic distance between pairs was then associated with genetic distance and plotted. BA.1* was excluded from the geographic distance versus genetic distance analysis because it showed too little geographic structure to be informative about viral spread, perhaps because of its rapid, nearly clonal rise or an unusually large number of introductions (**Fig S16**). Given we observed geographic structure for genomes differing by up to 12 substitutions, while more distantly related viruses were approximately uniformly distributed throughout the state, we can take this as the amount of divergence required for pairs to be distributed randomly throughout the state. As a simple model to translate this into time, we can treat the two viruses being compared as descending from a recent common ancestor, with each descendant lineage accumulating ∼0.5 mutations per generation. Thus a difference of 12 substitutions would accumulate in ∼12 generations or, assuming ∼5 days per generation, ∼60 days.

### Identification of closely related pairs

The identification of closely related cases was performed in Julia (v1.9). To identify all pairs of closely related cases within our dataset, first a random case was selected and all cases occurring within 10 days of this case were identified. Then the genetic distance from the initial randomly selected case to all of these temporally linked cases was calculated, and cases with a genetic distance of 2 or fewer SNPs were recorded as being closely related. This process was repeated until all cases had been compared against all other temporally linked cases. Resulting cases which had any number of additional closely related cases were then recorded as being “linked” while cases with no additional closely related cases were “unlinked.”

To compute the percent enrichment of closely related viruses within the same testing facility compared to within the same municipality, we calculated, per virus, the percent of contemporary (occurring within 10 days) same-facility cases which were closely related and subtracted the percent of contemporary (occurring within 10 days) same-municipality cases collected in public testing which were closely related. To additionally get a relative percent change, the likelihood of close relatives in each facility was calculated on a sector level (that is, summing the number of related cases for all cases within a given sector to avoid dividing by zero) and then divided by the percent of contemporary closely related in public testing in the community. Due to high variability in the likelihood of close relatives among public testing for specific age groups in each sector, we only calculated this on average for sectors, not within specific ages in each sector.

We then averaged all cases per sector to report summary findings. To describe these patterns over age groups we computed overlapping, 3-year rolling averages per sector. Age groups which had fewer than 25 viruses for comparison were dropped from the analysis.

### Detection of putative transmission events

An individual was defined as a transmitter when an index and contact met the following criteria: 1) the index and contact consensus sequences differed by no more than two consensus-level changes, 2) the consensus level changes in the contact were detected at sub-consensus levels in the index, 3) the contact appeared downstream of the index in a divergence phylogeny, 4) the collection date of the index was ≤6 days before that of the contact, and 5) if two genomes were identical, the defining mutations were at ≤90% frequency in the index, only seen <5 times in the community, and rose to 100% frequency in the contact.

### Estimation of sequencing rates to detect novel lineages

A multinomial regression model was applied to estimate the daily frequencies of variants BA.1*, BA.2*, BA.2.12.1*, BA.5*, and BQ.1*. Using the estimated variant frequencies, a simple model was used to estimate the earliest possible detection dates. This model incorporates a binomial draw, accounting for both sampling rates and the estimated frequencies of each variant, outputting the number of variant genomes that would be detected on a given day. This model was used to calculate the probability of detecting a variant genome by running it 1,000 times per genome sequencing rate (10, 50, 100, 200, up to 1,000 genomes per week). The resulting probabilities were used to estimate a 95% credible interval of the first date of detection for each variant under each sequencing rate.

We additionally applied the vartrack_prob_detect_cont function from PhyloSamp^19^ to verify these modeling results. For this function we set the coefficient of detection ratio to 1 (i.e. assuming no difference in the probability of detection between lineages), the initial prevalence to 0.0001, and the percent of samples which generate high quality genomes to 100% (to reflect that the modelling approach we employed relied on the number of high quality genomes). This function was then run with logistic growth rates of 0.075, 0.125, 0.175, and 0.225 and sampling rates ranging from 7 genomes per week to 1000 genomes per week.

### Estimation of sequencing rates to detect growth of novel lineages

To estimate how many genomes would be needed and when growth of variants would be detected after reaching 3% frequency, the dataset was subsampled 100 times at rates of 10, 50, 100, 200, 300, and so on, up to 1,000 genomes per week. For each subsample, the date when the lineage reached 3% frequency was recorded. A logistic regression was generated starting from day 1 (after reaching 3% frequency) through 30 days, capturing the day when the growth rate became positive with 95% probability. The probability was plotted that a positive 95% growth rate could be detected within each sampling rate N days after the lineage reached 3% frequency.

### Statistical comparison of date when each lineage reached 50% frequency

A logistic growth model was used to analyze the growth trajectories of Omicron sublineages (BA.1*, BA.2*, BA.2.12.1*, and BA.5*) in each sector. For each sector, the logistic growth model was fit using nonlinear least squares (NLS) regression to estimate the parameters growth rate and lineage frequency. The model for each lineage was constrained to only consider data starting from the first day where the proportion of genomes belonging to the lineage exceeded 0. The fitted models were used to generate predicted frequency of each lineage over time.

The tsum.test() function within BSDA^38^ was used to compare the growth rates of the variants between sectors. Specifically, the significance was tested of the differences between sectors in the timing of when each variant reached 50% frequency. The tsum.test() function was used while accounting for standard errors and sample sizes. A 95% confidence level was applied to assess the significance of differences in the growth rates and 50% threshold timing.

## Statistical analysis

All statistical analyses were performed in R (The R Core team, 2017), python 3, or Julia (v1.9). Pairwise ranksum tests for comparisons of Ct values were performed using the ranksum function in scipy^39^. All multiple comparisons were corrected using multipletests in statsmodels^35^ using the Benjamini-Hochberg correction. All additional statistical details including sample numbers can be found in the figure legends.

## Supplemental information titles and legends

**Figure S1. Genomic signatures of repeated and mixed SARS-CoV-2 infection.** Within our dataset, we were able to link genomes back to an individual within 90 days of their first genome.

A) Number of mixed lineage infections. X-axis: lineage of the consensus genome. Y-axis: lineage detected at sub-consensus frequencies. B) Number of mutations separating genomes collected from the same individual vs time between their collection dates. Colors indicate the same (gray) or different (blue) Pango lineages for the two genomes. Dashed lines delimit candidate region for evolution in persistently infected individuals (>14 days and >2 mutations).

C) Histogram showing the number of genomes per individual among those with more than one genome in the dataset. D) Histogram of the number of days between sequential genomes from the same individual; the dotted line denotes the median interval (4 days). E) Kernel density plot relating time between sample collection to the number of mutations observed; samples belonging to the same lineage cluster below 2 mutations and within 14 days, consistent with a single infection. F) Monthly prevalence of individuals with detected mixed infections, overlaid on a multinomial regression of circulating lineage frequencies.

**Figure S2. Temporal and demographic distribution of sequenced genomes.** A–C) Temporal distribution of the number of genomes collected grouped by A) age, B) municipality type, and C) gender. (D–E) Number of genomes collected within each facility type by D) age and E) gender.

**Figure S3. Overview of testing sectors and sampling intensity.** Left, table summarizing the testing sectors included in the dataset, showing the total number of genomes and the number of unique facilities per sector. Right, weekly histogram of genomes collected from each testing sector; the orange line indicates the total number of unique facilities contributing data in each week.

**Figure S4.** Genomes collected from public testing by age, as a fraction of the Massachusetts census population of that age.

**Figure S5. Robustness of sector-specific relatedness to subsampling.** Data were randomly subsampled per sector such that the sample size was the same in each sector and then bootstrap samples of fold-enrichment (x-axis) of closely related viruses within a facility versus between facility and municipality were plotted; dashed line: testing sector mean.

**Figure S6. Per facility enrichment of closely related viruses.** Scatterplot of the average enrichment in closely related viruses per facility compared to the square root of the total number of contemporary viral pairs in each facility, which is proportional to the total number of genomes each facility contributed to the complete dataset.

**Figure S7. Age-specific likelihood of close viral relatedness across settings.** Likelihood of identifying close viral relatives among cases within the same facility (colored lines) and among cases tested at public facilities within the same municipality (grey), stratified by age and sector. Curves show 3-year rolling averages for age groups with more than 30 samples; shaded regions denote bootstrapped 95% confidence intervals.

**Figure S8. Age-specific relatedness within public testing facilities.** Likelihood of identifying close viral relatives within the same testing facility as a function of age among public testing sites. Curves show 3-year rolling averages for age groups with more than 30 samples; shaded regions represent bootstrapped 95% confidence intervals.

**Figure S9. Lineage growth dynamics in college-aged individuals.** Logistic regressions of four SARS-CoV-2 lineages prevalence among 18- to 22-year-olds in college testing settings (teal) and in public testing (dark blue), compared with all public testing data (grey). Estimated dates at which each lineage reached 50% frequency are shown below, with 95% confidence intervals where available; for BA.5*, insufficient data precluded estimation of a 95% confidence interval for the 50% date.

**Figure S10. Lineage growth dynamics across testing settings.** Logistic regression fits and estimated dates at which lineage frequency reached 50% (with 95% confidence intervals) for BA.1, BA.2, BA.2.12.1, and BA.5, shown from top left to bottom right. Results are stratified by testing setting: public testing (blue), colleges (yellow), hospitals and clinics (red), and schools, employers, and skilled nursing facilities (green).

**Figure S11. Viral introductions scale with municipality population size and density across lineages.** Datasets were restricted to genomes collected prior to the date at which each major lineage reached 50% frequency in Massachusetts and were further subsampled to sequence 1% of confirmed infections within each geographic level (country, state, municipality) to account for urban–rural sampling biases. A) Scatter plots of the number of detected introductions per municipality as a function of municipality population, shown separately for each lineage; solid lines indicate linear regression fits with shaded 95% confidence intervals. B) Scatter plots of the number of detected introductions per municipality as a function of municipality population density, shown separately for each lineage; solid lines indicate linear regression fits with shaded 95% confidence intervals. C) Scatter plots of the number of detected introductions per municipality as a function of municipality population (left) and population density (right), averaged across all lineages; solid lines indicate linear regression fits with shaded 95% confidence intervals. Blue lines show the regression fits including Boston. Gray lines show regression fits removing Boston. The positive trends remain after removing Boston, although their strength is reduced.

**Figure S12. Vaccination uptake and infection timing in children.** A) Cumulative percentage of individuals aged 5–11 years receiving their first (black) and second (grey) COVID-19 vaccine doses following vaccine availability. B) Biweekly histograms showing the proportion of the Massachusetts population infected with Delta (top) and BA.1* (bottom) over time, stratified by age.

**Figure S13. Transmission patterns by demographic and clinical features.** A) Proportion of males and females who initiated at least one transmission event. B) Proportion of males and females who were linked versus unlinked. C) Proportion of individuals initiating a transmission event stratified by symptom status; symptom data were restricted to reports prior to January 1, 2022, and no significant difference was observed (p = 0.1). D) Proportion of symptomatic and asymptomatic individuals who were linked versus unlinked. E) Proportion of individuals initiating a transmission event by vaccination timing among vaccinated individuals; “early” vaccination corresponds to Feb 1 - May 1, 2021, while “late” vaccination corresponds to vaccination after booster availability (from Nov, 2021 - Jan 2023). F) Proportion of individuals initiating a transmission event by age group.

**Figure S14. Factors associated with SARS-CoV-2 viral load (Ct).** A) Heatmap of pairwise comparisons of lineage-specific Ct values; colored cells indicate significant differences after multiple-testing correction. The asterisk denotes the lineage label (e.g., BA.1*) and includes all sublineages following that designation, according to WHO/Nextstrain nomenclature. B) Temporal trends in mean lineage Ct values, showing lower Cts during lineage emergence and higher Cts during decline; shaded regions denote 95% confidence intervals. C) Violin plot comparing Ct values in symptomatic and asymptomatic cases. D) Ct values decrease with increasing time since last vaccination (−0.002 Ct per day; p < 2 × 10⁻¹⁶). E) Ct values decrease modestly with age (−0.001 Ct per year). F) Small but statistically significant differences in Ct by sex. G) Ct values increase with number of vaccine doses (+0.14 Ct per dose; p < 2 × 10⁻¹⁶).

**Figure S15. Sampling requirements for early lineage detection.** Application of PhyloSamp to estimate the number of high-quality genomes required to detect emerging lineages before they reach 1% prevalence. Curves are shown for growth rates comparable to BA.1 (fast-growing), BA.5 (slow-growing), and intermediate rates. The x-axis (plotted in reverse for comparability to Fig. 4A) shows the empirical probability of detecting a lineage before 1% prevalence, assuming logistic growth from an initial prevalence of 0.0001. The y-axis indicates the number of high-quality genomes generated per week.

**Figure S16. Reduced geographic structure during BA.1 emergence.** Relationship between genetic and geographic distance, shown as the density of the fraction of sequence pairs at a given SNP difference, during the exponential phase of BA.1* emergence (top; cases detected before December 21, 2021) and during the subsequent plateau and decline phase (bottom).

## Notes

### Competing Interest Statement

The authors have declared no competing interest.

### Author Declarations

The research project (Protocol #1603078) was reviewed and approved by the Massachusetts Department of Public Health (MADPH) Institutional Review Board and covered by a reliance agreement at the Broad Institute. An additional non-human subjects research determination was made by the Harvard Longwood Campus Institutional Review Board.

### Summary of Updates

Uploaded a corrected version of figure 2

